# Joint Heat and PM2.5 Exposure Across US Metropolitan Areas: Multi-Stressor Disparities, Historical Redlining, and a Multi-Metric Assessment Framework

**DOI:** 10.64898/2026.08.20.26360970

**Authors:** S. V. Mandalapu, R. Sharma, A. Pillarisetti

## Abstract

Many urban health outcomes are shaped by environmental stressors that occur together rather than in isolation, yet methods for measuring such co-occurrence at the neighbourhood scale remain underdeveloped. We developed a multi-metric framework for joint co-exposure assessment and applied it to characterise the joint spatial distribution of summer surface heat and fine particulate matter (PM2.5) across 42,304 census tracts in 48 large US metropolitan areas during summers 2015 to 2020, covering approximately 174.6 million residents. The framework combines a composite co-exposure index, a joint exceedance indicator, a conditional exceedance ratio that compares observed joint occurrence to within-group statistical independence, and an upper tail dependence parameter estimated using both the non-parametric Capéraà-Fougères-Genest estimator and a Gumbel copula, with bias-corrected and accelerated (BCa) confidence intervals obtained from a 5,000-replicate metropolitan-area block bootstrap. Among residents of predominantly Black tracts, 13.21% lived in neighbourhoods that simultaneously exceeded the within-metropolitan-area 80th percentile for both heat and PM2.5, compared with 3.33% of residents of predominantly White tracts; the corresponding heat-only and PM2.5-only ratios were 2.88 and 2.48. Residents of Home Owners Loan Corporation grade D tracts had 3.97 times the odds (95% confidence interval 2.79 to 5.66) of joint hotspot residence compared with grade A residents after adjustment for contemporary tract racial composition, poverty, renter-occupancy, and pre-1960 housing. The within-group conditional exceedance ratio at the 80th percentile was 2.29 in predominantly White tracts (95% BCa CI 1.81 to 2.78), 1.27 in predominantly Black tracts (0.71 to 1.56), and 1.13 in predominantly Hispanic tracts (0.70 to 1.41); the White interval excluded one while the Black and Hispanic intervals included one, which we interpret as power-limited given fewer contributing CBSAs. Magnitudes attenuated under near-surface air temperature surfaces but the direction and statistical significance of the primary findings were preserved. The framework is portable to other compound-exposure questions and supports cumulative-impact assessment.

## 1. Introduction

Urban environmental stressors rarely act alone. Heat, air pollution, noise, and limited green space tend to cluster in particular neighbourhoods and shape health through cumulative and, at times, compound processes (Berberian et al., 2022; Cushing et al., 2015; Tessum et al., 2021). The same urban features that elevate surface temperature—impervious cover, sparse tree canopy, and proximity to high-traffic infrastructure, among others—also concentrate air pollutants near the ground (Benz and Burney, 2021; Hsu et al., 2021). The distribution of these features across the urban landscape is uneven, and their contemporary configuration reflects long-running patterns of inequitable investment, land use, and zoning. Methods that quantify the joint spatial distribution of multiple stressors are needed if regulatory cumulative-impact tools and place-based interventions are to act on the configurations of burden that residents actually experience.

Three methodological gaps motivate this work. First, we uniquely, to our knowledge, have assessed whether the joint distribution of heat and PM2.5 produces neighbourhood disparities that exceed those produced by either stressor alone and whether such joint patterns persist in historically redlined neighbourhoods after adjustment for contemporary covariates. Single-city work suggests such patterns exist (Ahn, 2024) but does not establish whether the pattern holds across metropolitan areas. Second, the analytical metrics used to characterise co-occurrence are not typically unified into a coherent framework. A given joint exceedance rate can reflect independent accumulation of two marginal disadvantages or active spatial co-location above statistical independence; and the two have different implications for intervention (AghaKouchak et al., 2014; Renard and Lang, 2007). Third, there has been a lack of systematic testing of whether multi-stressor disparity findings are robust to choices about exposure surface, threshold, metric, or spatial modelling.

We develop a multi-metric framework for joint co-exposure assessment at the census tract level and demonstrate its use for summer surface heat and PM2.5 across 48 large US metropolitan areas. The framework includes four complementary metrics: a composite continuous index, a binary joint exceedance indicator, a conditional exceedance ratio that compares observed joint occurrence to within-group statistical independence, and an upper tail dependence parameter estimated using both non-parametric and parametric methods. We apply the framework to characterise the joint exposure distribution by predominant racial composition of the tract and by historical Home Owners Loan Corporation (HOLC) grading, a documented driver of contemporary neighbourhood-scale environmental disparity (Hoffman et al., 2020; Lane et al., 2022; Nowak et al., 2022; Shkembi and Neitzel, 2025; Swope et al., 2022). The application is presented as a demonstrative case for the framework; the primary contribution is methodological, the framework is implementable using publicly available exposure surfaces, and we provide reproducible code. We test the robustness of all reported associations across alternative exposure surfaces, exceedance thresholds, pollutant substitutions, study years, and spatial-dependence specifications, and report E-values to characterise robustness to unmeasured confounding.

## 2. Methods

### 2.1 Study design

We conducted a cross-sectional ecological analysis at the census tract level. The analytic sample comprised 42,304 census tracts in 48 large US metropolitan areas (Core Based Statistical Areas, CBSAs) during summers (June through August) 2015 to 2020, covering approximately 174.6 million residents (Table S10). We included 48 of the 49 CBSAs covered by our exposure data sources; San Juan-Bayamón-Caguas, PR, was excluded because the EPA Fused Air Quality Surface Downscaler (Section 2.2) does not cover Puerto Rico. Tract demographic data were derived from the American Community Survey 2016 to 2020 five-year estimates, aligned with the exposure window.

### 2.2 Exposure assessment

Tract summer heat exposure was characterised using daytime land surface temperature (LST) from the MODIS MOD11A1 Collection 6.1 product, extracted via Google Earth Engine with quality filtering. Tract summer mean PM2.5 was obtained from the US EPA Fused Air Quality Surface Downscaler (FAQSD), which combines ground-monitor data with chemistry-transport model output (Berrocal et al., 2012). Ground-level ozone was obtained from the same FAQSD product for sensitivity analyses. For cross-source sensitivity we used two alternative PM2.5 surfaces (Di et al., 2019; Atmospheric Composition Analysis Group V5 satellite-derived) and two alternative temperature surfaces (PRISM, Daymet).

### 2.3 Joint exposure metrics

Four complementary metrics were constructed, each capturing a distinct dimension of joint burden.

#### Composite Co-Exposure Index

Within each CBSA, summer mean LST and summer mean PM2.5 were standardised to z-scores, and the composite co-exposure index (CCEI) was defined as their arithmetic mean (Eq. 1), where Z_LST,i and Z_PM2.5,i denote the within-CBSA z-scores for tract i. Equal weighting was adopted as the parsimonious default; principal-component validation and 30/70 and 70/30 weight sensitivities are reported in Section 3.7.

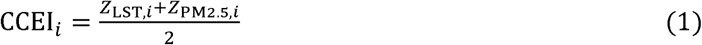

#### Joint exceedance

Tracts simultaneously exceeding the within-CBSA 80th percentile of both Z_LST and Z_PM2.5 were classified as joint hotspots (Eq. 2): where q80 is the 80th-percentile threshold and I(·) is the indicator function. The 80th percentile was chosen a priori as a conservative cumulative-impact threshold; sensitivities across the 70th to 95th percentile are reported in Section 3.7.

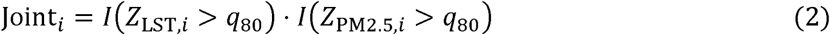

#### Conditional exceedance ratio

The joint exceedance indicator alone does not distinguish whether elevated joint occurrence reflects independent accumulation of two marginal disadvantages or active spatial co-clustering above what marginal rates would predict. For each predominant-racial-group subsample we computed the conditional exceedance ratio (CER) as the ratio of the observed joint exceedance rate to the rate expected under statistical independence of the two stressors within the group (Eq. 3), where P_g denotes the within-group population-weighted probability and p = 0.80 in the primary specification. A CER above one indicates within-group spatial co-clustering above statistical independence; a CER of one indicates within-group independence.

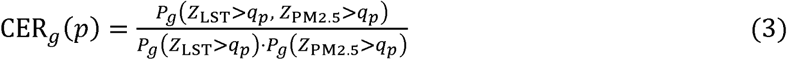

#### Upper tail dependence

Strength of co-occurrence at exposure extremes was characterised by the upper tail dependence coefficient lambda_U. We report the empirical conditional exceedance function chi(u) = P(Z_PM2.5 > q_u | Z_LST > q_u) by group across thresholds u in (0.70, 0.975) as the primary model-free summary of joint extreme behaviour. As a parametric and semi-parametric cross-check, we also estimated lambda_U using the Capéraà-Fougères-Genest (CFG) estimator (Capéraà et al., 1997), which is non-parametric within the extreme-value copula family, and using a Gumbel copula (Eqs. 4 and 5). The Gumbel family was selected as the parametric reference because it parameterises asymmetric upper-tail dependence among standard Archimedean families (Nelsen, 2006); we present CFG and Gumbel estimates in parallel and treat their agreement as cross-method validation within the extreme-value class. The empirical chi(u) curve is the assumption-free quantity. All tail dependence metrics characterise spatial co-occurrence of long-term mean exposures across tracts within a metropolitan area; they do not characterise temporal co-occurrence of episodic compound events at fixed locations.

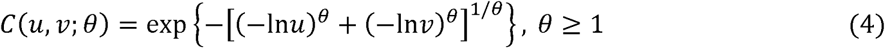

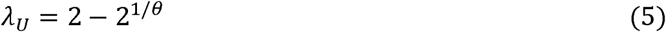

### 2.4 Demographic, historical, and health data

Tract racial composition was derived from ACS 2016 to 2020 five-year estimates. Tracts were classified by predominant racial composition using a 50% threshold; we treat these as descriptive subgroup labels for population-weighted comparison. Historical HOLC redlining grades from the Mapping Inequality project (Nelson et al., 2020) were crosswalked from HOLC neighbourhood polygons to 2020 census tracts; HOLC coverage applies to 12,200 tracts (28.8% of analytic sample) in 43 CBSAs. Tract prevalence estimates for selected chronic conditions were obtained from CDC PLACES (Centers for Disease Control and Prevention, 2025); we note that PLACES estimates are model-based small-area projections rather than direct measurements, and we use them descriptively only.

### 2.5 Statistical analysis

Disparities were quantified as population-weighted ratios of exposure or joint hotspot prevalence across predominant-racial-composition subsamples. Multilevel linear, logistic, and Poisson models with tracts nested within CBSAs (Bates et al., 2015) were fitted with sequential adjustment for tract poverty, renter-occupancy, and pre-1960 housing share (Models 1 through 4) and a random slope on tract percent people of colour (Model 5). Cluster-robust standard errors at the CBSA level used the Bell-McCaffrey CR2 correction (Pustejovsky and Tipton, 2018). The fully adjusted Model 4 specification for the CCEI outcome is given in Eq. 6, where POC_ij, POV_ij, RENT_ij, and PRE1960_ij are the standardised tract-level shares of people of colour, residents below the federal poverty line, renter-occupied housing units, and housing units built before 1960; u_j is a CBSA random intercept; and ε_ij is a tract-level residual.

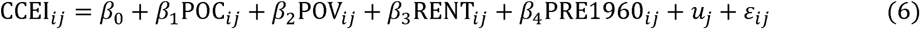

For HOLC-graded tracts we estimated multilevel logistic regressions of joint hotspot status on HOLC grade with sequential adjustment for the same contemporary covariates (M0 unadjusted through M4 fully adjusted), with a CBSA random intercept. We refer to the M0-to-M4 progression as a sequential adjustment or attenuation analysis rather than as causal mediation, because the no-unmeasured-mediator-confounding assumption required for formal natural-effect decomposition cannot be established for a 1930s exposure. Region-stratified adjusted odds ratios were computed within Northeast, Midwest, and South Census regions; the Western region contained too few HOLC grade A tracts with non-zero joint hotspot prevalence to support stable estimation in the random-intercept logistic specification.

Confidence intervals for the conditional exceedance ratio, the chi(u) curve, and the copula tail dependence parameters were obtained from a CBSA-block bootstrap that resamples CBSAs with replacement and retains all tracts within each resampled CBSA, respecting within-CBSA spatial dependence. For the conditional exceedance ratio we report bias-corrected and accelerated (BCa) intervals (Efron, 1987), computed from 5,000 bootstrap replicates with the acceleration parameter estimated from a leave-one-CBSA-out cluster jackknife using the same CER estimator (Eq. 3). The 80th-percentile thresholds defining LST and PM2.5 exceedance were computed once on the full 48-CBSA analytic sample within each CBSA and held fixed across all bootstrap replicates and jackknife folds; under this design the BCa intervals are anchored on the full-sample point estimates of Eq. 3, which are reproducible from the marginals in Table 2. For other tail dependence quantities we report percentile intervals from 1,000 bootstrap replicates, which are well-centred on their estimators in this design. Pairwise group contrasts in tail dependence parameters were computed as the proportion of bootstrap replicates yielding a non-positive difference. We assessed robustness across six sensitivity dimensions: alternative PM2.5 and temperature surfaces; exceedance thresholds from the 70th to 95th percentile; CCEI weighting; ozone-for-PM2.5 substitution; study-year stratification; and stratification by region and CBSA size. As sensitivity to spatial dependence we estimated spatial autoregressive (SAR) lag models in the 10 largest CBSAs. To assess robustness to unmeasured confounding we computed E-values (VanderWeele and Ding, 2017) for the primary multilevel POC coefficient and for the HOLC grade D-vs-A comparison.

Analyses used R 4.5.2 with the data.table, lme4, sf, spdep, copula, EValue, and ggplot2 packages (Bates et al., 2015; Bivand and Wong, 2018; Hofert et al., 2018; Pebesma, 2018; R Core Team, 2025; Wickham, 2016). Reproducible code is available at a public repository (URL provided on the title page; withheld here for peer review).

## 3. Results

### 3.1 Sample description

The analytic sample comprised 42,304 census tracts in 48 large US metropolitan areas (Table 1). Across summers 2015 to 2020, tracts had a mean summer LST of 35.0 degrees C, a mean PM2.5 of 9.36 micrograms per cubic metre, and a mean 8-hour maximum ozone of 43.9 parts per billion. Approximately 48% of the study population identified as people of colour, with substantial heterogeneity across CBSAs (interquartile range 22 to 75%). The unconditional tract joint hotspot rate was 6.7%; 10.1 million residents lived in joint hotspot tracts.

**Table 1.**
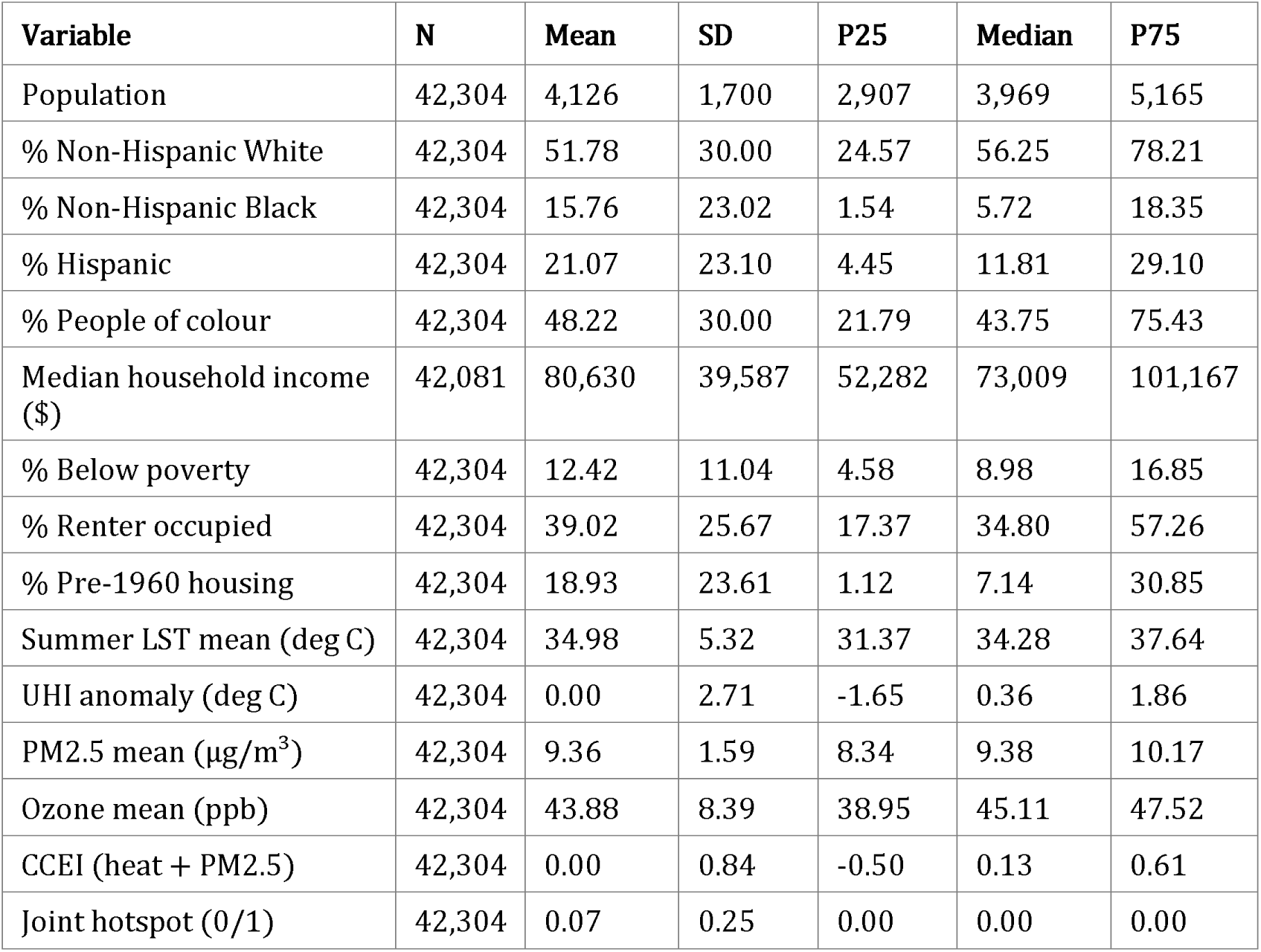
Descriptive statistics for the analytic sample (n = 42,304 census tracts in 48 metropolitan areas, summers 2015 to 2020).

### 3.2 Marginal and joint exposure by predominant racial composition

Single-stressor exposures differed across racial groups (Table 2). Population-weighted within-CBSA exceedance of the 80th percentile of LST occurred in 31.85% of residents of predominantly Black tracts compared with 11.06% of residents of predominantly White tracts (heat-only ratio 2.88); the corresponding PM2.5-only exceedance rates were 32.56% and 13.15% (PM2.5-only ratio 2.48). Joint exceedance of both thresholds occurred in 13.21% of residents of predominantly Black tracts compared with 3.33% of residents of predominantly White tracts, a 3.97-fold joint disparity that exceeded either single-stressor ratio (Fig. 1, Panels A-D).

**Table 2.** Population-weighted marginal and joint exposure rates by predominant racial composition.

| Group | Tracts (n) | P(LST>q80) (%) | P(PM>q80) (%) | P(joint) (%) | Heat ratio vs W | PM ratio vs W | Joint ratio vs W |
| --- | --- | --- | --- | --- | --- | --- | --- |
| White | 23,670 | 11.06 | 13.15 | 3.33 | 1.00 | 1.00 | 1.00 |
| Black | 4,306 | 31.85 | 32.56 | 13.21 | 2.88 | 2.48 | 3.97 |
| Hispanic | 5,605 | 33.43 | 26.07 | 9.88 | 3.02 | 1.98 | 2.97 |

*Note: Predominant racial composition defined as 50% or more of tract population. Within-CBSA thresholds at the 80th percentile. Rates are population-weighted using ACS 2016 to 2020 tract total population. Mixed (no single 50%+ group, 8,003 tracts) and predominantly Asian tracts (720) are reported in Supplementary Table S9.*

**Figure 1.**
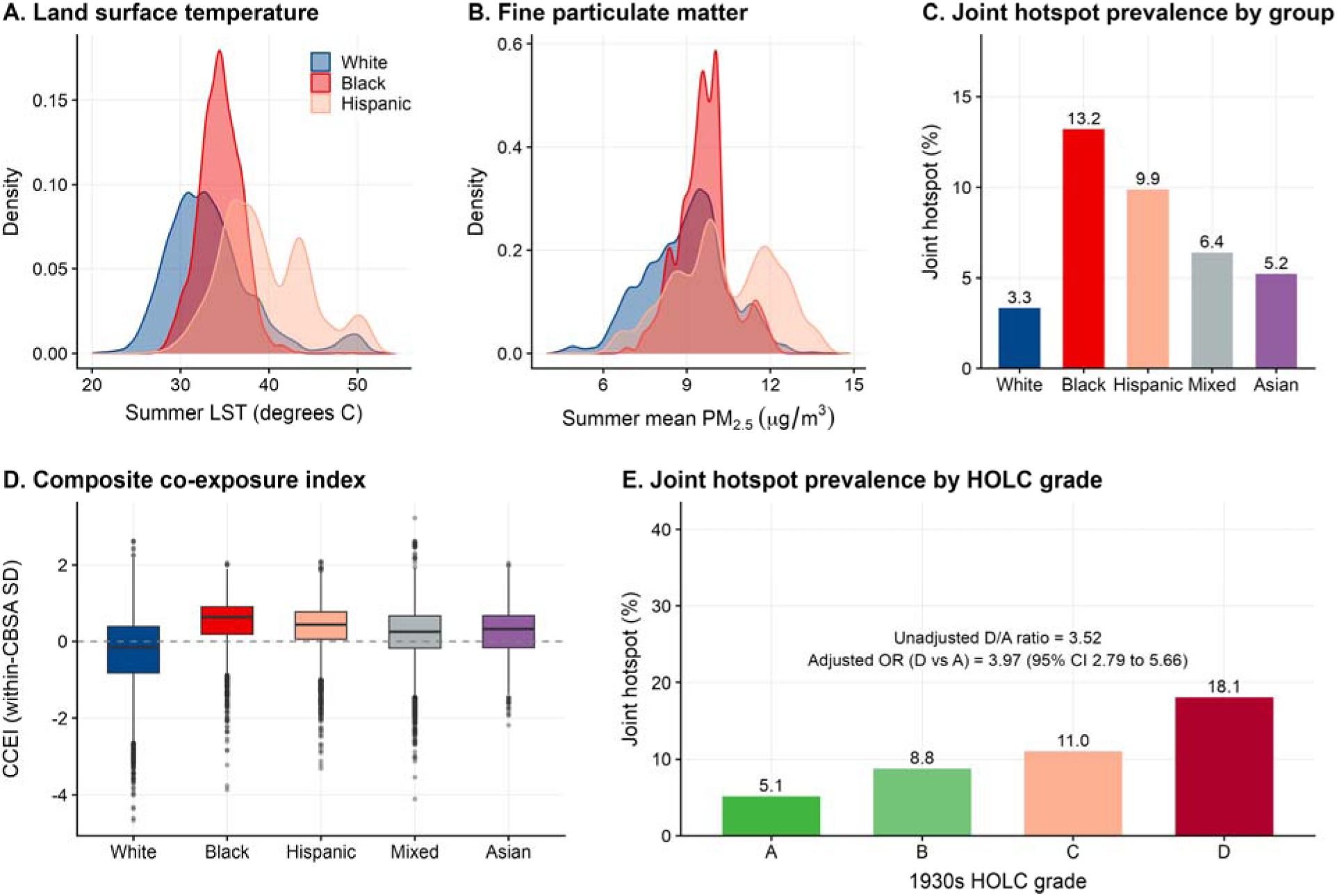
Joint heat and PM2.5 exposure by predominant racial composition and HOLC grade. Panel A: tract-level distribution of summer mean land surface temperature (degrees C) by predominant racial group. Panel B: tract-level distribution of summer mean PM2.5 (micrograms per cubic metre) by predominant racial group. Panel C: population-weighted joint hotspot rate (%) by predominant racial group, with the within-CBSA 80th-percentile threshold definition. Panel D: composite co-exposure index (CCEI, z-score) by predominant racial group as boxplots. Panel E: population-weighted joint hotspot rate (%) by HOLC grade (A, B, C, D), annotated with the unadjusted D-vs-A ratio of 3.52 and the fully adjusted Model 4 odds ratio of 3.97 (95% CI 2.79 to 5.66). Sample: 42,304 census tracts across 48 metropolitan areas, summers 2015 to 2020; HOLC panel restricted to 12,200 HOLC-graded tracts in 43 metropolitan areas.

It is worth setting the observed joint disparity against the value that would obtain under statistical independence of the two stressors within each group. The independence-implied within-group joint exceedance rate is the product of the two within-group marginal exceedance rates. Under independence, the predominantly Black within-group joint rate would be 10.37% (31.85% × 32.56%) and the predominantly White within-group joint rate would be 1.45% (11.06% × 13.15%), giving an independence-implied Black-White joint disparity of 7.15. The observed Black-White joint disparity of 3.97 is therefore substantially below the independence-implied value. The reason is within-group co-clustering, quantified by the conditional exceedance ratio (Section 3.5), which is larger in predominantly White tracts than in predominantly Black or Hispanic tracts. The observed joint disparity is large in absolute terms and exceeds either single-stressor disparity; it does not reflect a within-group multiplicative amplification of one stressor by the other.

### 3.3 Joint exposure in historically redlined tracts

Joint exposure was concentrated in tracts graded D by the 1930s Home Owners Loan Corporation (Table 5, Panel A). Population-weighted joint hotspot prevalence was 5.14% in grade A tracts, 8.78% in grade B, 11.01% in grade C, and 18.10% in grade D, a monotonic gradient with an unadjusted D/A ratio of 3.52. Mean CCEI was 0.10 in grade A and 0.52 in grade D. Contemporary tract characteristics differed sharply by HOLC grade: grade D tracts contained on average 26.4% non-Hispanic Black residents and 21.4% below the federal poverty line, compared with 12.1% and 8.6% in grade A tracts (Fig. 1, Panel E).

In the fully adjusted multilevel logistic model (M4), residents of grade D tracts had 3.97 times the odds (95% confidence interval 2.79 to 5.66) of joint hotspot residence compared with residents of grade A tracts after adjustment for contemporary tract racial composition, poverty, renter-occupancy, and pre-1960 housing share (Table 5, Panel B). Sequential adjustment from M0 (unadjusted) through M4 attenuated the odds ratio non-monotonically: from 5.94 unadjusted to 4.19 with racial composition, 3.87 with poverty, 3.49 with renter-occupancy, and then 3.97 with pre-1960 housing share. The non-monotonic step from M3 to M4 reflects that pre-1960 housing share acts as a suppressor in this specification: conditional on the M3 covariates, including pre-1960 housing in the model raises rather than lowers the HOLC contrast. The fully adjusted M4 odds ratio of 3.97 is attenuated from the unadjusted M0 value of 5.94 but remains substantially elevated, indicating that the four contemporary covariates measured here account for a notable but not majority fraction of the unadjusted association. We interpret the residual as the portion not captured by the four measured tract covariates, which is consistent with long-run differences in transportation siting, industrial land use, infrastructure investment, and tree canopy that contemporary demographic and housing variables capture imperfectly (Hoffman et al., 2020; Nowak et al., 2022; Shkembi and Neitzel, 2025; Swope et al., 2022).

Region-stratified analysis showed substantial geographic variation in the HOLC gradient (Table S6). The adjusted D-vs-A odds ratio was 7.85 (95% CI 3.77 to 16.31) in the Midwest, 5.45 (2.62 to 11.37) in the Northeast, and 1.82 (1.06 to 3.13) in the South. The Midwest and Northeast confidence intervals overlap heavily and the supportable claim is that the Southern HOLC gradient is weaker than the Northern and Midwestern gradients, rather than a clean three-region rank ordering; all three regional intervals exclude one. The Western odds ratio is not reported because the Western HOLC subsample produced a singular Hessian under the random-intercept logistic specification. The per-CBSA HOLC gradient was monotonically increasing from A to D in 18 of 43 HOLC-coverage CBSAs and weakly monotonic (D > A) in 27 of 43 (Table S7). The largest D-minus-A gaps occurred in Jacksonville (D rate 73.7%, A rate 0%), Philadelphia (57.8%, 0%), and Baltimore (53.3%, 0%); the 0% A rate in these three CBSAs reflects small A-grade tract counts (3, 45, and 20 tracts respectively) and the corresponding D/A ratios are not defined. A few CBSAs (Houston, New Orleans, San Antonio, Memphis, Providence, Charlotte) show negative D-minus-A gaps, but these reverse from small grade-A tract samples within those CBSAs and we read them as small-sample artefacts rather than substantive gradient reversals.

### 3.4 Multilevel models of the racial gradient

Multilevel models showed that the racial gradient in co-exposure persisted after adjustment for tract-level socioeconomic and built-environment characteristics (Table 3). In the fully adjusted Model 4 (n = 42,304 tracts in 48 CBSAs), a one standard deviation increase in percent people of colour was associated with a 0.366 standard deviation increase in the CCEI (cluster-robust standard error 0.024, p < 0.001). Pre-1960 housing share (β = 0.175) and renter-occupancy (β = 0.153) were independent positive predictors; tract poverty was a small negative independent predictor (β = −0.085) once racial composition and built-environment metrics were included, consistent with the known partial decoupling of poverty and racial composition in heat exposure (Hsu et al., 2021). The intra-class correlation of 0.055 indicated that approximately 95% of the variance in CCEI was within rather than between CBSAs.

**Table 3.** Multilevel linear models of CCEI, with sequential adjustment for tract covariates (M1 through M4) and random slope on percent people of colour (M5).

| Predictor | M1 | M2 | M3 | M4 | M5 |
| --- | --- | --- | --- | --- | --- |
| Intercept | 0.070 | 0.067 | 0.072 | 0.085 | 0.108 |
| % People of colour | 0.440 | 0.425 | 0.379 | 0.366 | 0.385 |
| % Below poverty | n/a | 0.025 | -0.059 | -0.085 | -0.084 |
| % Renter occupied | n/a | n/a | 0.178 | 0.153 | 0.150 |
| % Pre-1960 housing | n/a | n/a | n/a | 0.175 | 0.172 |
| ICC (CBSA) | 0.069 | 0.065 | 0.065 | 0.055 | 0.130 |
| AIC | 94,888 | 94,866 | 93,446 | 91,856 | 91,236 |
| N tracts | 42,304 | 42,304 | 42,304 | 42,304 | 42,304 |
| N CBSAs | 48 | 48 | 48 | 48 | 48 |

*Note: All continuous predictors standardised. Coefficients are linear-mixed-model fixed effects on the CCEI (a within-CBSA z-scored composite). Cluster-robust standard errors use the Bell-McCaffrey CR2 correction.*

In multilevel logistic and Poisson models for joint hotspot status (Table 4), a one standard deviation increase in percent people of colour was associated with an odds ratio of 1.75 (95% CI 1.66 to 1.86) and a rate ratio of 1.62 (95% CI 1.54 to 1.70). Pre-1960 housing share (odds ratio 1.77) and renter-occupancy (1.52) were positive independent predictors; the tract poverty coefficient was statistically null in the logistic model once the other tract covariates were included.

**Table 4.** Multilevel logistic and Poisson regression of joint hotspot status on tract-level covariates.

| Predictor | Logistic OR (95% CI) | Poisson RR (95% CI) |
| --- | --- | --- |
| Intercept | 0.05 (0.04, 0.06) | 0.05 (0.04, 0.06) |
| % People of colour | 1.75 (1.66, 1.86) | 1.62 (1.54, 1.70) |
| % Below poverty | 1.04 (0.99, 1.09) | 1.02 (0.98, 1.06) |
| % Renter occupied | 1.52 (1.44, 1.60) | 1.45 (1.39, 1.53) |
| % Pre-1960 housing | 1.77 (1.69, 1.85) | 1.61 (1.55, 1.68) |

*Note: Joint hotspot = within-CBSA exceedance of the 80th percentile for both LST and PM2.5. All predictors standardised. Models include a CBSA random intercept; cluster-robust SEs via CR2 correction.*

**Table 5.** Joint co-exposure by HOLC grade. Panel A: descriptive statistics by grade. Panel B: sequential adjustment of the HOLC D-vs-A association.

| **HOLC grade** | **Tracts (n)** | **CCEI** | **Joint (%)** | **% NH Black** | **% Poverty** | **Joint rate ratio**<br>**(vs A)** |
| --- | --- | --- | --- | --- | --- | --- |
| A | 745 | 0.10 | 5.14 | 12.1 | 8.6 | 1.00 |
| B | 2,410 | 0.35 | 8.78 | 18.9 | 13.2 | 1.71 |
| C | 5,512 | 0.46 | 11.01 | 20.4 | 17.1 | 2.14 |
| D | 3,533 | 0.52 | 18.10 | 26.4 | 21.4 | 3.52 |

Panel B. Sequential adjustment of the HOLC D-vs-A association.
| Model | OR D vs A | 95% CI | % of unadjusted log-OR attenuated |
| --- | --- | --- | --- |
| M0: HOLC only (unadjusted) | 5.94 | (4.25, 8.31) | 0.0 |
| M1: + tract % POC | 4.19 | (2.97, 5.90) | 19.6 |
| M2: + poverty | 3.87 | (2.74, 5.47) | 24.0 |
| M3: + renter occupancy | 3.49 | (2.46, 4.94) | 29.9 |
| M4: + pre-1960 housing | 3.97 | (2.79, 5.66) | 22.6 |

Note: HOLC sample restricted to 12,200 tracts in 43 metropolitan areas with HOLC coverage. Multilevel logistic regressions of joint hotspot status with a CBSA random intercept. Attenuation = $(\log_{OR\_M0} - \log_{OR\_Mk}) / \log_{OR\_M0}$ . Pre-1960 housing acts as a suppressor at the final step (M3 to M4), and the M4 odds ratio is attenuated from but remains substantially elevated relative to the unadjusted M0 estimate.

Multilevel model residuals exhibited strong spatial autocorrelation (global Moran’s I above 0.83 across CBSAs, all p < 0.001). Spatial autoregressive lag models in the 10 largest CBSAs attenuated the within-tract POC coefficient substantially, with the mean coefficient declining from 0.366 in Model 4 to approximately 0.04. We interpret the multilevel Model 4 coefficient as a population-average gradient that absorbs spatial co-clustering of tract-level demographic and environmental features, and the SAR estimate as a stricter conditional-association estimate that is, by construction, harder to identify in spatially structured data. The descriptive joint disparity reported in Section 3.2, the conditional exceedance ratios reported in Section 3.5, and the HOLC findings reported in Section 3.3 are computed outside the multilevel-regression framework and are unaffected by the SAR specification.

### 3.5 Within-group spatial co-clustering and tail dependence

The within-group conditional exceedance ratio at the 80th percentile, computed from the full-sample population-weighted marginal and joint exceedance rates in Table 2, was 2.29 in predominantly White tracts (95% BCa CI 1.81 to 2.78), 1.27 in predominantly Black tracts (0.71 to 1.56), and 1.13 in predominantly Hispanic tracts (0.70 to 1.41) (Table S5). Point estimates exceed one in all three groups, indicating positive within-group co-clustering on average. The 95% BCa interval excludes one for the White group; the Black and Hispanic intervals include one. BCa intervals were computed from a 5,000-replicate CBSA-block bootstrap with acceleration estimated from a leave-one-CBSA-out cluster jackknife (Section 2.5); they are constructed around the full-sample point estimates and reproducible from the joint and marginal exceedance rates in Table 2.

The number of CBSAs contributing to each group’s CER80 estimate differs substantially (Table S2). Predominantly White tracts are present in all 48 CBSAs (median 400 tracts per CBSA, range 77 to 2,128). Predominantly Black tracts are present in 38 CBSAs (median 58, range 1 to 608), with 23 contributing 50 or more tracts. Predominantly Hispanic tracts are present in 40 CBSAs (median 34, range 1 to 1,270), with 17 contributing 50 or more tracts. The smaller and more variable block counts for the Black and Hispanic groups produce wider bootstrap confidence intervals; we interpret the inclusion of one in those intervals as power-limited rather than as positive evidence for marginal independence.

The empirical conditional exceedance function chi(u) = P(Z_PM2.5 > q_u | Z_LST > q_u) is shown by group across thresholds u from 0.70 to 0.975 (Fig. 2, Table S8). At moderate thresholds (u between 0.70 and 0.90), point estimates of chi(u) for predominantly White and predominantly Black tracts are higher than for predominantly Hispanic tracts; predominantly White and Black point estimates are similar, with overlapping confidence intervals at every u value. At the most extreme threshold tested (u = 0.975), the point-estimate ordering shows predominantly Black tracts above predominantly White tracts (0.148, 95% CI 0.019 to 0.261 versus 0.057, 0.019 to 0.120); the confidence intervals overlap at u = 0.975. We present chi(u) as the model-free quantity of primary interest. As parametric and semi-parametric cross-checks we report CFG non-parametric and Gumbel parametric estimates of upper tail dependence in Table S3 and Figure 3 (CFG: White 0.301, Black 0.243, Hispanic 0.099; Gumbel: 0.307, 0.250, 0.068). CFG and Gumbel agree within 0.06 across all three groups, indicating that the parametric assumption of the Gumbel family does not drive the tail dependence findings within the extreme-value-copula class. The Hispanic-versus-others contrast is statistically robust across both estimators (CFG White-minus-Hispanic 0.203, bootstrap p = 0.005; CFG Black-minus-Hispanic 0.144, p = 0.018); the White-versus-Black contrast is not statistically distinguishable under the block bootstrap (CFG difference 0.058, p = 0.246).

**Figure 2.**
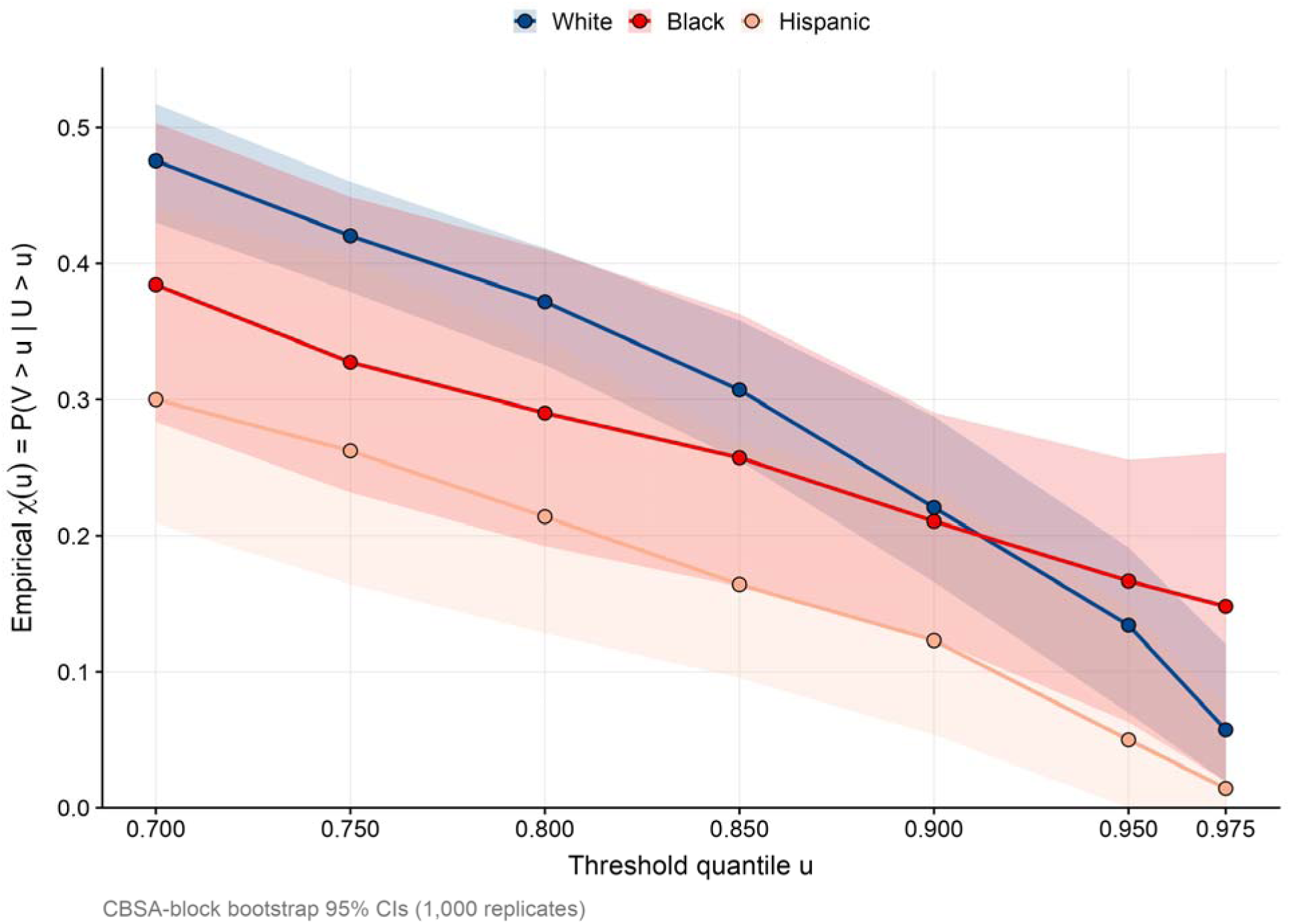
Empirical conditional exceedance function chi(u) = P(Z_PM2.5 > q_u | Z_LST > q_u) by predominant racial composition across thresholds u from 0.70 to 0.975. Shaded ribbons indicate 95% confidence intervals from CBSA-block bootstrap (1,000 replicates). Sample: 23,670 predominantly White tracts, 4,306 predominantly Black tracts, 5,605 predominantly Hispanic tracts.

**Figure 3.**
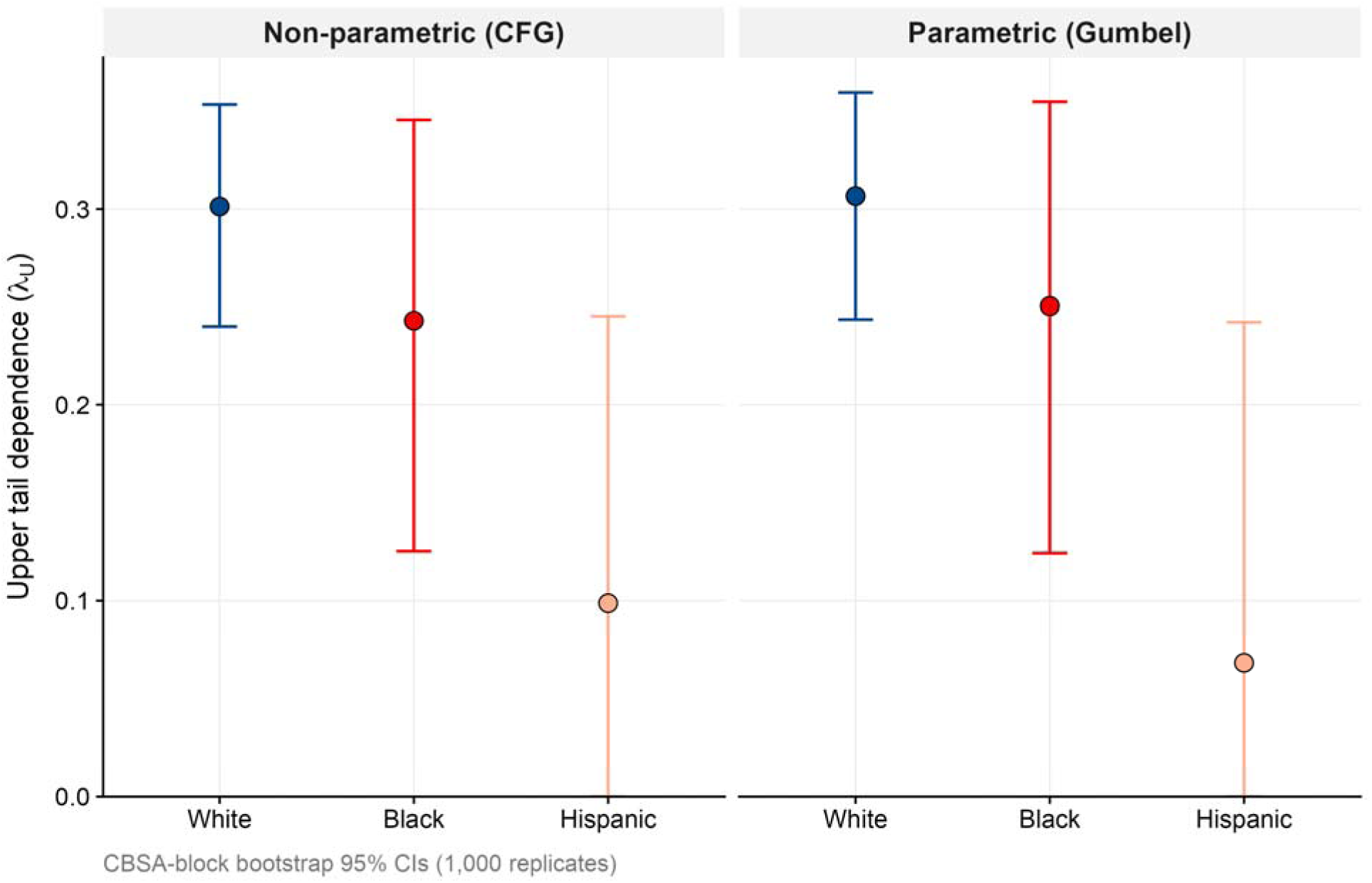
Race-stratified upper tail dependence (lambda_U). Panel A: Caperaa-Fougeres-Genest non-parametric estimator by predominant racial group with CBSA-block bootstrap 95% confidence intervals. Panel B: Gumbel copula parametric estimator by predominant racial group with CBSA-block bootstrap 95% confidence intervals. Agreement between estimators within 0.06 across racial groups provides cross-method validation within the extreme-value-copula class. Sample as Figure 2.

### 3.6 Geographic variation

The racial gradient in co-exposure varied across metropolitan areas (Fig. 4). The largest CBSA-level Black-White CCEI gaps occurred in Minneapolis-St. Paul (random-effect-adjusted POC slope 0.66), Jacksonville (0.66), Pittsburgh (0.55), Cincinnati (0.54), and Tampa (0.53); the flattest gradients occurred in New Orleans (0.06), Buffalo (0.18), San Diego (0.18), and Phoenix (0.18). Region-stratified Black-White joint hotspot ratios were 4.74 in the Midwest, 3.94 in the South, and 3.06 in the Northeast. The Western region contained no tracts where Black residents constituted 50% or more of the population, so the categorical Black-White joint ratio is not estimable for Western CBSAs; the continuous percent people of colour gradient remained positive and large in Western CBSAs (β = 0.416 in the multilevel Model 4 within-Western subsample). CBSA-level Black-White dissimilarity correlated modestly with the CBSA-level Black-White CCEI gap (Pearson r = 0.33, p = 0.022), and a cross-level interaction between tract percent people of colour and CBSA dissimilarity was small but statistically significant (β = 0.0075, p = 0.001), indicating modestly steeper within-tract racial gradients in more segregated metropolitan areas.

**Figure 4.**
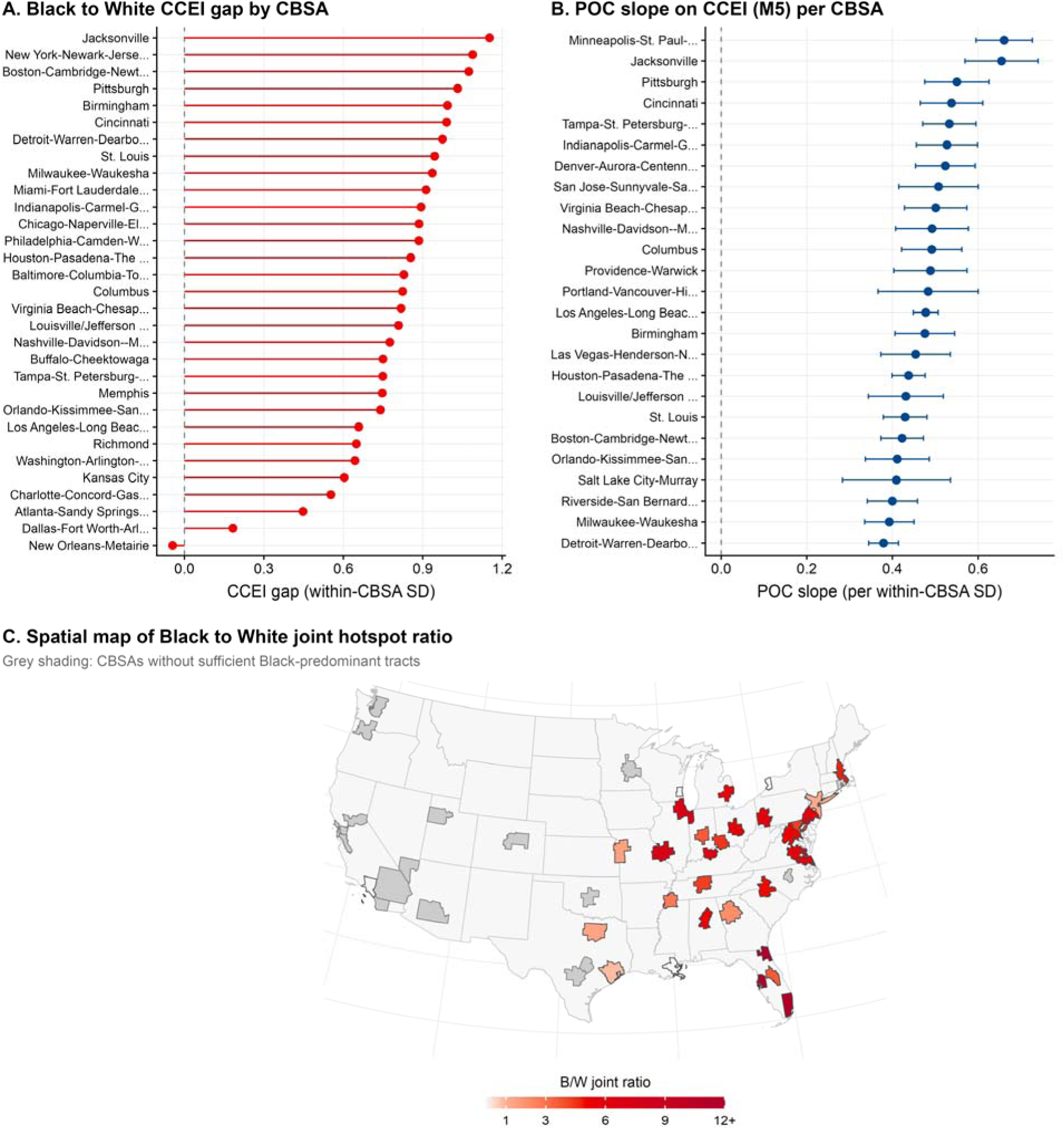
Geographic variation in the racial gradient of joint co-exposure across CBSAs. Panel A: CBSA-level Black-White CCEI gap (standard deviation units), ordered from largest to smallest. Panel B: random-effect-adjusted slope on tract percent people of colour from Model 5, with 95% confidence intervals, by CBSA. Panel C: spatial map of population-weighted Black-White joint hotspot ratio by CBSA, with hatching for CBSAs lacking tracts with 50% or more Black residents. Sample: 48 metropolitan areas, summers 2015 to 2020.

### 3.7 Sensitivity and robustness

Disparity conclusions were robust in direction and statistical significance across six sensitivity dimensions, although magnitudes varied (Fig. 5, Tables 6 and S1). The Black-White joint hotspot ratio remained between 3.39 and 4.36 across exceedance thresholds from the 70th to 95th percentile (Table 6). Replacing PM2.5 with ground-level ozone preserved direction and statistical significance with a smaller magnitude (Black-White joint ratio 2.9). The percent people of colour coefficient varied by less than 4% across study years. The equal-weighted CCEI correlated with the first principal component of standardised LST and PM2.5 at r = 0.968, supporting the equal-weighting choice; the racial gradient was qualitatively unchanged under 30/70 and 70/30 weighting.

**Figure 5.**
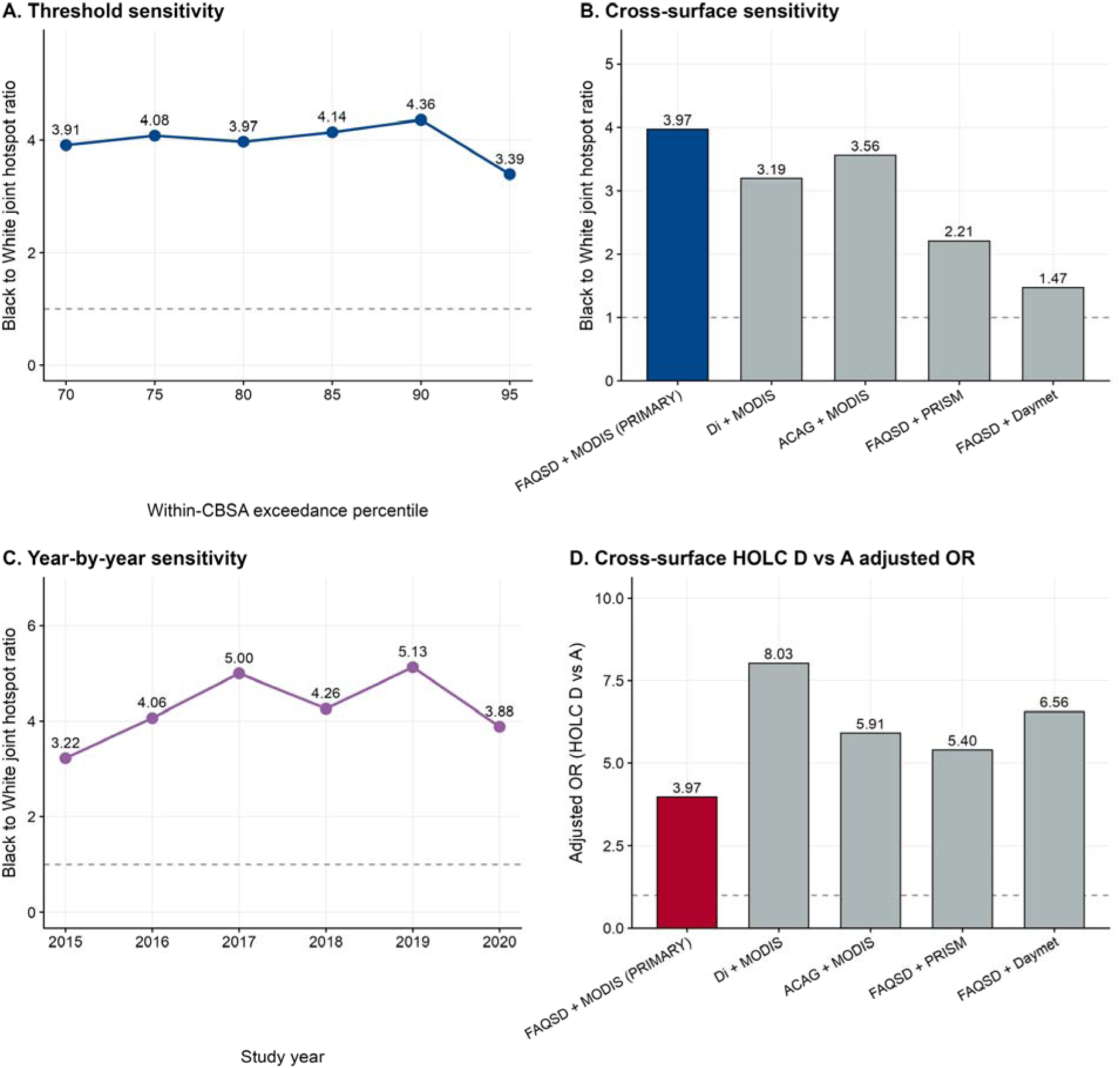
Sensitivity analyses summary. Panel A: Black-White joint hotspot ratio across exceedance thresholds from the 70th to 95th percentile. Panel B: Black-White joint hotspot ratio across the five exposure-surface combinations. Panel C: Black-White joint hotspot ratio across the six study years (2015 to 2020). Panel D: HOLC D-vs-A adjusted odds ratio across the five exposure-surface combinations, demonstrating that the primary FAQSD-plus-MODIS specification gives the most conservative HOLC estimate. All panels include 95% confidence intervals where computable.

**Table 6.** Threshold sensitivity of the Black-White joint hotspot disparity.

| Threshold | Black (%) | White (%) | Gap (pp) | Ratio |
| --- | --- | --- | --- | --- |
| 70th percentile | 28.11 | 7.19 | 20.92 | 3.91 |
| 75th percentile | 20.46 | 5.02 | 15.44 | 4.08 |
| 80th percentile (primary) | 13.21 | 3.33 | 9.88 | 3.97 |
| 85th percentile | 8.29 | 2.00 | 6.29 | 4.14 |
| 90th percentile | 4.27 | 0.98 | 3.29 | 4.36 |
| 95th percentile | 1.22 | 0.36 | 0.86 | 3.39 |

Note: Joint hotspot = simultaneous within-CBSA exceedance of the threshold-percentile for both LST and PM2.5. Population-weighted rates by predominant racial composition (50% threshold). Disparity ratio remains between 3.39 and 4.36 across the 70th to 95th percentile.

Cross-source sensitivity using two alternative PM2.5 products and two alternative temperature products preserved the direction and statistical significance of disparities across all five surface combinations, but magnitudes attenuated substantially under near-surface air temperature surfaces. The Black-White joint hotspot ratio was 3.97 under the primary FAQSD+MODIS specification, 3.19 under Di+MODIS, 3.56 under ACAG+MODIS, 2.21 under FAQSD+PRISM, and 1.47 under FAQSD+Daymet (Table S1). The attenuation under PRISM and Daymet is consistent with the established observation that satellite-retrieved LST exaggerates spatial contrasts in impervious-surface-heavy urban tracts relative to near-surface air temperature (Hsu et al., 2021). The HOLC D-vs-A adjusted odds ratio was substantially larger under every alternative surface (range 5.40 to 8.03) than under the primary FAQSD+MODIS specification (3.97), indicating that the primary specification provides the most conservative HOLC estimate among the surfaces examined.

E-values indicated that an unmeasured confounder with an approximate risk-ratio association of 2.14 (lower confidence bound 2.11) with both the exposure and the outcome conditional on measured covariates would be required to fully explain away the primary multilevel POC coefficient, and an approximate risk-ratio of 7.41 (lower bound 5.03) to fully explain away the HOLC grade D-vs-A association (Table S4). The E-value calibrates robustness of the population-average multilevel coefficient; it does not characterise robustness of the spatial-conditional estimate from the SAR specification, which attenuates substantially under explicit spatial structure as noted in Section 3.4. All primary hypothesis tests survived both Benjamini-Hochberg and Bonferroni correction.

## 4. Discussion

We develop a multi-metric framework for joint co-exposure assessment and demonstrate its use for summer surface heat and PM2.5 across 48 large US metropolitan areas. Three substantive findings emerge from the application. First, the joint Black-White disparity in heat and PM2.5 exceedance (3.97) exceeds either single-stressor disparity (heat 2.88, PM2.5 2.48), so single-stressor cumulative-impact analyses understate the joint burden borne by residents of predominantly non-White tracts. Second, joint exposure is associated with historical Home Owners Loan Corporation grading: residents of grade D tracts had 3.97 times the odds of joint hotspot residence compared with residents of grade A tracts after adjustment for contemporary tract covariates, and the four measured contemporary covariates account for only a modest fraction of the unadjusted gradient. Third, within-group co-clustering above statistical independence is statistically distinguishable from independence in predominantly White tracts; point estimates for predominantly Black and Hispanic tracts are similarly elevated but our current sample, with fewer contributing CBSAs for those groups, cannot statistically distinguish them from independence. These findings are descriptive cross-sectional associations; the analysis does not establish causal pathways from historical HOLC grading or contemporary demographic composition to joint exposure.

### 4.1 Historical redlining and contemporary co-exposure

The HOLC findings reported here are consistent with a body of work documenting associations between the 1930s grades and contemporary environmental and health outcomes (Hoffman et al., 2020; Huang and Sehgal, 2022; Lane et al., 2022; Nowak et al., 2022; Shkembi and Neitzel, 2025; Swope et al., 2022). Two aspects of our findings extend prior work. First, the joint co-exposure outcome captures a dimension of cumulative burden that single-stressor redlining studies do not measure; the unadjusted D/A joint hotspot ratio of 3.52 is steeper than published heat-only or PM2.5-only HOLC gradients of similar order. Second, the sequential adjustment analysis describes the fraction of the unadjusted HOLC gradient that is consistent with contemporary tract characteristics. Across the M0-to-M4 progression, racial composition is associated with the largest single attenuation, poverty and renter-occupancy with smaller marginal attenuations, and pre-1960 housing share acts as a suppressor at the final step (conditional on the M3 covariates, including pre-1960 housing raises rather than lowers the HOLC contrast). The fully adjusted M4 odds ratio of 3.97 is attenuated from the unadjusted 5.94 but remains substantially elevated, indicating that the contemporary tract covariates measured here account for a notable but not majority fraction of the historical-to-present association. We are explicit that this is a descriptive cross-sectional finding: the analysis does not establish causal pathways from 1930s grading to contemporary co-exposure. The regional pattern is consistent with the historical geography of the HOLC programme, which operated most extensively in older Northeastern and Midwestern industrial cities. The contemporary HOLC association in our data is correspondingly strongest in those two regions and weaker in the South. We do not have sufficient grade A coverage in Western CBSAs to estimate the Western regional association.

### 4.2 Joint disparity and within-group co-clustering

The conditional exceedance ratio sharpens the interpretation of the joint disparity. A naive reading of the 3.97 joint Black-White ratio might assume that heat and PM2.5 multiplicatively co-cluster within communities of colour. The CER analysis indicates that this reading is not supported by the data. The CER at the 80th percentile is 2.29 in predominantly White tracts (95% BCa CI 1.81 to 2.78), 1.27 in predominantly Black tracts (0.71 to 1.56), and 1.13 in predominantly Hispanic tracts (0.70 to 1.41). The White interval excludes one; the Black and Hispanic intervals include one. As noted in Section 3.5 the estimates draw on fewer CBSA blocks than the White estimate, and we read the inclusion of one in those CIs as power-limited rather than as evidence for marginal independence. The joint burden borne by residents of predominantly non-White tracts is large in absolute terms and reflects the simultaneous occurrence of two elevated marginal disadvantages; whether there is additional within-group co-clustering above what those marginal rates would predict is a question that future work with more statistical power should resolve.

### 4.3 Methodological considerations for joint-burden assessment

The multi-metric framework applied here lends itself to cumulative-impact screening and is portable across compound-exposure questions in urban health. The four metrics answer related but distinct questions. The composite index characterises the continuous gradient. The joint exceedance indicator identifies acutely affected neighbourhoods. The conditional exceedance ratio quantifies whether elevated joint occurrence reflects spatial co-clustering above marginal independence. The empirical chi(u) curve characterises the conditional probability of one extreme given another and is model-free. The CFG and Gumbel estimators provide parametric and semi-parametric cross-checks within the extreme-value-copula family; CFG is non-parametric within that class rather than assumption-free, and we recommend that the empirical chi(u) curve be reported alongside any copula-derived tail dependence summary.

Two methodological choices warrant comment. First, confidence intervals for the CER, chi(u), and tail dependence parameters were obtained from a CBSA-block bootstrap rather than a tract-level bootstrap. Tract-level resampling treats tracts as exchangeable draws, incompatible with the strong within-CBSA spatial autocorrelation in the data. The CBSA-block bootstrap respects within-metropolitan-area dependence by resampling CBSAs as the independent units. It produces wider but more honest confidence intervals; our finding that the within-group White-versus-Black tail dependence contrast is not statistically distinguishable under the block bootstrap is a consequence. Second, the tail dependence and CER metrics characterise spatial co-occurrence of long-term mean exposures across tracts within a metropolitan area; they do not characterise temporal co-occurrence of episodic compound heat-pollution events at a fixed location, which would require daily-resolution exposure data linked to individual health outcomes (Schnell and Prather, 2017). The spatial framework we present is suited to the structural-determinants question that this paper addresses; the temporal framework remains a target for future linked-data work.

### 4.4 Implications for cumulative-impact assessment

Single-stressor exposure assessments dominate the regulatory landscape under EJScreen and CalEnviroScreen. Our findings argue for the routine integration of joint-burden metrics into such tools, including a within-CBSA joint exceedance indicator and a historical structural indicator such as HOLC grade where available. The joint disparity reported here (3.97) is larger than either single-stressor marginal disparity (2.88 for heat, 2.48 for PM2.5); regulatory analyses that report only marginal disparities therefore understate the joint burden in a direction-correct sense, although the joint and marginal disparities are computed on different outcomes and a precise quantitative percentage of understatement is not well defined. The HOLC E-value of 7.41 is approximately three times the magnitude of the population-of-colour E-value (2.14), indicating that the redlining gradient in this analysis is unusually robust to plausible unmeasured confounding, with the spatial-conditional caveat noted in Section 3.4.

Joint hotspot tracts had modestly higher prevalence of several CDC PLACES outcomes than non-hotspot tracts. The joint hotspot population in this study is approximately 10.1 million; applying tract-level prevalence differences yields illustrative ecological projections of order 110,000 additional diabetes cases and 165,000 additional cases of frequent poor mental health. We emphasise that these are descriptive ecological projections, not individual-level attributions: the cross-sectional design does not support causal inference, individual-level risk factors (smoking, body mass, healthcare access, occupational exposures) are not measured, and PLACES estimates are model-based small-area projections that incorporate demographic and socioeconomic covariates in their prediction algorithms.

### 4.5 Limitations

MODIS land surface temperature is a remote-sensing proxy for ambient air temperature and does not capture indoor exposure or humidity-dependent heat stress. Cross-source sensitivity (Table S1) shows that the Black-White joint disparity attenuates from 3.97 (primary FAQSD+MODIS specification, computed on the full analytic sample) to 1.47 (FAQSD+Daymet) when LST is replaced with near-surface air temperature; the direction and statistical significance of the disparity are preserved across all five surface combinations, but magnitudes are not. Future work should compare LST-derived disparities against disparities derived from in-situ air temperature observations where dense networks exist. The HOLC analysis covers 28.8% of the analytic sample and concentrates in older Eastern and Midwestern CBSAs in line with the geography of the HOLC programme; the HOLC findings may not generalise to Sun Belt or Western CBSAs that developed after the redlining era. The 50% predominance threshold for the categorical Black-White comparison is not estimable in Western CBSAs because no Western tract in our sample exceeds 50% Black residents; the continuous percent people of colour gradient remains positive and large in Western CBSAs. Multilevel residuals exhibited strong spatial autocorrelation, and the within-tract POC coefficient attenuates substantially under spatial autoregressive specification; the descriptive joint disparity, the conditional exceedance ratios, and the HOLC findings are computed outside the multilevel framework and are unaffected. CDC PLACES estimates are model-based small-area projections; the ecological health-burden projections reported in Section 4.4 are illustrative, not causal. The framework characterises spatial co-occurrence of long-term mean exposures, not temporal co-occurrence of episodic compound events.

## 5. Conclusions

Across 48 large US metropolitan areas, summer surface heat and PM2.5 exceedances are associated in particular neighbourhoods. The Black-White joint disparity in our analytic sample (3.97) exceeds either single-stressor disparity, joint exposure is associated with historical Home Owners Loan Corporation grade D status (adjusted odds ratio 3.97, 95% CI 2.79 to 5.66), and within-group co-clustering above statistical independence is statistically distinguishable from independence in predominantly White tracts but underpowered to be distinguished in the Black and Hispanic groups. These are descriptive cross-sectional associations and do not establish causal pathways. Single-stressor cumulative-impact screening understates joint burden, and the multi-metric framework described here, with metropolitan-area block-bootstrap inference, is portable to other compound-exposure questions and is implementable using publicly available exposure surfaces. Reproducible analysis code is provided to support adoption.

## Data Availability

All data used in this study are publicly available; data sources are documented in the Methods section of the manuscript.

## CRediT author contributions

S.V.M.: Conceptualisation, Methodology, Software, Formal analysis, Data curation, Investigation, Validation, Writing - Original Draft, Writing - Review and Editing, Visualisation. R.S.: Investigation, Methodology, Validation, Writing - Review and Editing. A.P.: Investigation, Methodology, Validation, Writing - Review and Editing, Project administration, Resources, Supervision.

## Funding

This research received no specific grant from any funding agency in the public, commercial, or not-for-profit sectors.

## Competing interests

The authors declare no competing interests.

## Ethics approval

Not applicable. This study uses publicly available, de-identified aggregate data at the census tract level and was therefore exempt from institutional review board review.

## Data and code availability

All data used in this study are publicly available; data sources are documented in the Methods section of the manuscript. Reproducible analysis code is available at https://github.com/saivenkatmandalapu/co-exposure-penalty

## Notes

### Competing Interest Statement

The authors have declared no competing interest.

