## supplementary material for "Joint Heat and PM2.5 Exposure Across US Metropolitan Areas: Multi-Stressor Disparities, Historical Redlining, and a Multi-Metric Assessment Framework"

**Contents**

This document contains ten supplementary tables (S1 to S10) and additional methodological notes referenced in the main manuscript. Tables are organised in the order of their first citation in the main text.

**Table S1.** Cross-source sensitivity of disparity findings across five exposure-surface combinations.

**Table S2.** Tail dependence analysis sample sizes by predominant racial group.

**Table S3.** Race-stratified upper tail dependence with CBSA-block bootstrap confidence intervals.

**Table S4.** E-values for the primary multilevel coefficient and the HOLC grade D versus A comparison.

**Table S5.** Within-group conditional exceedance ratio at the 80th percentile, full-sample point estimates with BCa intervals.

**Table S6.** Region-stratified HOLC grade D versus A adjusted odds ratios.

**Table S7.** Per-CBSA HOLC grade A and D joint hotspot rates (43 HOLC-coverage CBSAs).

**Table S8.** Empirical conditional exceedance function chi(u) by predominant racial group across thresholds u.

**Table S9.** Population-weighted marginal and joint exposure rates for tracts of mixed and predominantly Asian racial composition.

**Table S10.** The 48 metropolitan areas in the analytic sample.

**Table S1. Cross-source sensitivity**

*Cross-source sensitivity of disparity findings across five exposure-surface combinations. The Black-White joint hotspot ratio and the HOLC D versus A adjusted odds ratio are reported under the primary FAQSD+MODIS specification and four alternative PM2.5 and temperature surfaces.*

| **Surface combination** | **B-W joint hotspot ratio** | **B-W CCEI gap (SD)** | **HOLC D vs A adjusted OR** | **N tracts** |
| --- | --- | --- | --- | --- |
| FAQSD + MODIS (primary) | 3.97 | 0.71 | 3.97 | 42,304 |
| Di et al. + MODIS | 3.19 | 0.62 | 8.03 | 42,302 |
| ACAG + MODIS | 3.56 | 0.78 | 5.91 | 42,244 |
| FAQSD + PRISM | 2.21 | 0.48 | 5.40 | 42,301 |
| FAQSD + Daymet | 1.47 | 0.52 | 6.56 | 42,264 |

*Note: Each row is computed on the complete-case subsample for that surface combination; sample sizes vary by less than 0.2% across rows (range 42,244 to 42,304 tracts), so sample composition differences are likely not the source of magnitude variation across surfaces. All entries are within-CBSA z-score standardised. The HOLC D-vs-A adjusted OR is from a multilevel logistic model adjusting for tract percent people of colour, poverty, renter-occupancy, and pre-1960 housing, with a CBSA random intercept. The three PM2.5 products examined are FAQSD, Di et al. (2019), and the Atmospheric Composition Analysis Group V5 satellite-derived surface; the three temperature products are MODIS LST, PRISM, and Daymet.*

**Table S2. Tail dependence sample sizes**

*Tail dependence analysis sample sizes by predominant racial group, with population totals and per-CBSA tract distribution.*

| **Group** | **Tracts (n)** | **Population (M)** | **CBSAs contributing** | **Median tracts per CBSA** | **CBSAs with ≥50 tracts** |
| --- | --- | --- | --- | --- | --- |
| White | 23,670 | 96.5 | 48 | 400 | 48 |
| Black | 4,306 | 14.9 | 38 | 58 | 23 |
| Hispanic | 5,605 | 24.9 | 40 | 34 | 17 |

*Note: Predominant racial composition defined as 50% or more of tract population. CBSAs contributing are those with at least one tract in the group. The smaller and more variable block counts for the Black and Hispanic groups produce wider bootstrap confidence intervals; see Section 3.5 of the main manuscript for discussion.*

**Table S3. Race-stratified upper tail dependence**

*Race-stratified upper tail dependence parameters with 95% CBSA-block bootstrap percentile confidence intervals from 1,000 replicates. CFG is the non-parametric Capéraà-Fougères-Genest estimator (non-parametric within the extreme-value-copula family). Gumbel is the parametric Gumbel copula estimator. CER80 is the within-group conditional exceedance ratio at the 80th percentile.*

| **Group** | **CFG λU (95% CI)** | **Gumbel λU (95% CI)** | **CER80 (95% BCa CI)** |
| --- | --- | --- | --- |
| White | 0.301 (0.240, 0.353) | 0.307 (0.244, 0.359) | 2.29 (1.81, 2.78) |
| Black | 0.243 (0.125, 0.345) | 0.250 (0.124, 0.355) | 1.27 (0.71, 1.56) |
| Hispanic | 0.099 (0.000, 0.245) | 0.068 (0.000, 0.242) | 1.13 (0.70, 1.41) |

*Note: CFG and Gumbel intervals are 1,000-replicate percentile intervals. CER80 intervals are 5,000-replicate BCa intervals anchored on the full-sample CER80 point estimates from Table 2 marginals; see Section 2.5 of the main manuscript for the BCa procedure.*

**Table S4. E-values**

*E-values for the primary multilevel coefficient on percent people of colour and the HOLC grade D versus A comparison. The E-value is the minimum approximate risk-ratio strength of association with both the exposure and the outcome (conditional on measured covariates) that an unmeasured confounder would need to fully explain away the observed association.*

| **Comparison** | **Beta or log-OR** | **Approximate RR** | **E-value (estimate)** | **E-value (lower CI)** |
| --- | --- | --- | --- | --- |
| POC → CCEI (M4 multilevel) | 0.366 | 1.40 | 2.14 | 2.11 |
| HOLC D vs A → joint hotspot (M4 adjusted) | 1.380 | 3.97 | 7.41 | 5.03 |

*Note: E-values computed per VanderWeele and Ding (2017), as E = RR + sqrt(RR × (RR - 1)) for the point-estimate version, with the lower-CI version computed using the lower 95% confidence bound of the risk-ratio. The HOLC E-value of 7.41 indicates that an unmeasured confounder with approximate risk-ratio associations of 7.41 with both HOLC grade D residence and joint hotspot status (conditional on measured covariates) would be required to fully explain away the observed association. The E-value calibrates the population-average multilevel coefficient; it does not characterise robustness of the spatial-conditional estimate from the SAR specification.*

**Table S5. Conditional exceedance ratio (CER80)**

*Within-group conditional exceedance ratio at the 80th percentile. Point estimates are computed from the full-sample population-weighted marginal and joint exceedance rates in Table 2 of the main manuscript. The 95% BCa confidence intervals are from a 5,000-replicate CBSA-block bootstrap with acceleration estimated from a leave-one-CBSA-out cluster jackknife, with within-CBSA thresholds fixed at the full-sample 80th-percentile values.*

| **Group** | **Observed joint (%)** | **Independence-implied joint (%)** | **CER80 (95% BCa CI)** | **Excludes 1.0?** |
| --- | --- | --- | --- | --- |
| White | 3.33 | 1.45 | 2.29 (1.81, 2.78) | yes |
| Black | 13.21 | 10.37 | 1.27 (0.71, 1.56) | no |
| Hispanic | 9.88 | 8.72 | 1.13 (0.70, 1.41) | no |

*Note: Independence-implied joint rate is the product of the within-group marginal exceedance rates from Table 2. Worked example for White: 3.33 / (11.06% × 13.15%) = 3.33 / 1.45 = 2.29. The White interval excludes one. The Black and Hispanic intervals include one; these are interpreted as power-limited given fewer contributing CBSAs (Table S2) rather than as positive evidence for marginal independence.*

**Table S6. Region-stratified HOLC analysis**

*Region-stratified HOLC grade D versus A adjusted odds ratios for joint hotspot residence, from multilevel logistic models adjusting for tract racial composition, poverty, renter-occupancy, and pre-1960 housing, with a CBSA random intercept.*

| **Region** | **Tracts (n)** | **n grade A** | **n grade D** | **Adjusted OR (D vs A)** | **95% CI** |
| --- | --- | --- | --- | --- | --- |
| All HOLC-coverage | 12,200 | 745 | 3,533 | 3.97 | (2.79, 5.66) |
| Northeast | 4,335 | 196 | 1,193 | 5.45 | (2.62, 11.37) |
| Midwest | 2,916 | 151 | 848 | 7.85 | (3.77, 16.31) |
| South | 2,178 | 218 | 727 | 1.82 | (1.06, 3.13) |
| West | 2,771 | 180 | 765 | not estimated | not estimated |

*Note: The Western regional adjusted OR is not reported because the Western HOLC subsample produced a singular Hessian under the random-intercept logistic specification. This is most likely quasi-complete separation: Western grade A tracts have very low joint hotspot prevalence, and the within-Western CBSA count with both grade A and grade D representation is small, leaving the D-vs-A contrast weakly identified within a random-intercept structure. A Firth-corrected fit was not pursued because the West contributes little to the cross-region story. Tract counts match Table 5 in the main manuscript. The Northeast and Midwest 95% confidence intervals overlap heavily; the supportable claim is that the Southern HOLC gradient is weaker than the Northern and Midwestern gradients, rather than a clean three-region rank ordering; all three estimable regional intervals exclude one. All models are multilevel logistic regressions with HOLC grade as a four-level factor (A reference), tract racial composition, poverty, renter-occupancy, and pre-1960 housing as additional standardised covariates, and a CBSA random intercept; the OR reported is for the D-vs-A contrast.*

**Table S7. Per-CBSA HOLC analysis**

*Per-CBSA HOLC grade A and D joint hotspot rates and unadjusted D-minus-A gap (percentage points). Restricted to the 43 metropolitan areas with HOLC coverage in the analytic sample, sorted by D-minus-A gap (descending).*

| **CBSA** | **n tracts** | **Grade A rate (%)** | **Grade D rate (%)** | **D minus A (pp)** |
| --- | --- | --- | --- | --- |
| Jacksonville, FL | 47 | 0.0 | 73.7 | 73.7 |
| Philadelphia-Camden-Wilmington | 506 | 0.0 | 57.8 | 57.8 |
| Baltimore-Columbia-Towson | 276 | 0.0 | 53.3 | 53.3 |
| Columbus, OH | 123 | 7.7 | 58.3 | 50.6 |
| Atlanta-Sandy Springs-Roswell | 171 | 0.0 | 50.0 | 50.0 |
| St. Louis, MO-IL | 216 | 12.5 | 57.9 | 45.4 |
| Boston-Cambridge-Newton | 508 | 0.0 | 43.3 | 43.3 |
| Richmond, VA | 77 | 14.3 | 57.1 | 42.9 |
| Tampa-St. Petersburg-Clearwater | 133 | 0.0 | 41.4 | 41.4 |
| Indianapolis-Carmel-Greenwood | 179 | 7.7 | 45.1 | 37.4 |
| Louisville/Jefferson County | 91 | 0.0 | 37.0 | 37.0 |
| Minneapolis-St. Paul-Bloomington | 226 | 4.3 | 41.2 | 36.8 |
| Nashville-Davidson | 73 | 9.1 | 40.9 | 31.8 |
| Denver-Aurora-Centennial | 125 | 0.0 | 27.3 | 27.3 |
| Birmingham, AL | 88 | 25.0 | 51.9 | 26.9 |
| Chicago-Naperville-Elgin | 1,179 | 0.0 | 23.0 | 23.0 |
| Detroit-Warren-Dearborn | 580 | 12.5 | 33.3 | 20.8 |
| Miami-Fort Lauderdale | 196 | 0.0 | 16.4 | 16.4 |
| Kansas City, MO-KS | 170 | 0.0 | 13.8 | 13.8 |
| Los Angeles-Long Beach-Anaheim | 1,574 | 0.0 | 11.0 | 11.0 |
| Milwaukee-Waukesha | 260 | 0.0 | 10.1 | 10.1 |
| Phoenix-Mesa-Chandler | 34 | 0.0 | 10.0 | 10.0 |
| Oklahoma City, OK | 74 | 26.7 | 35.0 | 8.3 |
| Virginia Beach-Chesapeake-Norfolk | 120 | 0.0 | 6.0 | 6.0 |
| Seattle-Tacoma-Bellevue | 188 | 0.0 | 5.9 | 5.9 |
| New York-Newark-Jersey City | 2,938 | 0.0 | 2.1 | 2.1 |
| San Francisco-Oakland-Fremont | 392 | 0.0 | 0.7 | 0.7 |
| Sacramento-Roseville-Folsom | 51 | 0.0 | 0.0 | 0.0 |
| San Diego-Chula Vista-Carlsbad | 170 | 0.0 | 0.0 | 0.0 |
| San Jose-Sunnyvale-Santa Clara | 52 | 0.0 | 0.0 | 0.0 |
| Cincinnati, OH-KY-IN | 61 | 0.0 | 0.0 | 0.0 |
| Buffalo-Cheektowaga, NY | 148 | 0.0 | 0.0 | 0.0 |
| Portland-Vancouver-Hillsboro | 140 | 0.0 | 0.0 | 0.0 |
| Pittsburgh, PA | 273 | 27.8 | 27.8 | 0.0 |
| Dallas-Fort Worth-Arlington | 218 | 0.0 | 0.0 | 0.0 |
| Austin-Round Rock-San Marcos | 58 | 0.0 | 0.0 | 0.0 |
| Salt Lake City-Murray, UT | 59 | 0.0 | 0.0 | 0.0 |
| Houston-Pasadena-The Woodlands | 145 | 34.6 | 26.7 | -7.9 |
| New Orleans-Metairie, LA | 168 | 12.5 | 0.0 | -12.5 |
| San Antonio-New Braunfels, TX | 103 | 18.2 | 0.0 | -18.2 |
| Memphis, TN-MS-AR | 75 | 50.0 | 28.1 | -21.9 |
| Providence-Warwick, RI-MA | 114 | 54.5 | 28.6 | -26.0 |
| Charlotte-Concord-Gastonia, NC-SC | 46 | 100.0 | 47.4 | -52.6 |

*Note: The D-minus-A gap is computed as the percentage-point difference between grade D and grade A joint hotspot rates within the CBSA. CBSAs with 0% grade A rates (e.g., Jacksonville, Philadelphia, Baltimore) have undefined D/A ratios; these are typically CBSAs with very few grade A tracts (3, 45, and 20 tracts respectively in the three listed examples). The D > A pattern holds in 27 of 43 HOLC-coverage CBSAs.*

**Table S8. Empirical conditional exceedance function chi(u)**

*Empirical conditional exceedance function chi(u) = P(Z_PM2.5 > q_u | Z_LST > q_u) by predominant racial group across thresholds u from 0.70 to 0.975, with 95% percentile confidence intervals from a 1,000-replicate CBSA-block bootstrap.*

| **Group** | **u = 0.70** | **u = 0.75** | **u = 0.80** | **u = 0.85** | **u = 0.90** | **u = 0.95** | **u = 0.975** |
| --- | --- | --- | --- | --- | --- | --- | --- |
| White | 0.475 (0.430, 0.517) | 0.420 (0.379, 0.460) | 0.372 (0.326, 0.411) | 0.307 (0.255, 0.358) | 0.221 (0.166, 0.288) | 0.134 (0.070, 0.191) | 0.057 (0.019, 0.120) |
| Black | 0.384 (0.284, 0.503) | 0.328 (0.232, 0.449) | 0.290 (0.192, 0.410) | 0.257 (0.163, 0.363) | 0.211 (0.123, 0.290) | 0.167 (0.063, 0.256) | 0.148 (0.019, 0.261) |
| Hispanic | 0.300 (0.209, 0.442) | 0.262 (0.164, 0.403) | 0.214 (0.128, 0.344) | 0.164 (0.096, 0.270) | 0.123 (0.054, 0.236) | 0.050 (0.000, 0.148) | 0.014 (0.000, 0.076) |

*Note: chi(u) is reported as the model-free quantity of primary interest for tail dependence. The Hispanic-versus-others contrast is statistically robust at all thresholds (bootstrap p < 0.05); the White-versus-Black contrast is not statistically distinguishable at any threshold given overlapping confidence intervals. The point-estimate ordering at u = 0.975 shows Black exceeding White, but the confidence intervals overlap.*

**Table S9. Marginal and joint exposure rates for Mixed and Asian groups**

*Population-weighted marginal and joint exposure rates for tracts not in any predominant (50%+) Black, White, or Hispanic group (Mixed, 8,003 tracts) and for predominantly Asian tracts (720). Computed using the same within-CBSA 80th-percentile thresholds as Table 2 in the main manuscript (thresholds defined on the full 48-CBSA analytic sample, applied per group).*

| **Group** | **Tracts (n)** | **Population (M)** | **P(LST>q80) (%)** | **P(PM>q80) (%)** | **P(joint) (%)** | **Heat ratio vs W** | **PM ratio vs W** | **Joint ratio vs W** |
| --- | --- | --- | --- | --- | --- | --- | --- | --- |
| Mixed | 8,003 | 35.15 | 22.22 | 20.93 | 6.41 | 2.01 | 1.59 | 1.93 |
| Asian | 720 | 3.13 | 17.93 | 26.90 | 5.21 | 1.62 | 2.05 | 1.57 |

*Note: “P(LST>q80)” is the population-weighted within-group probability that summer LST exceeds the within-CBSA 80^th^ percentile; “P(PM>q80)” is the analogous probability for PM2.5; “P(joint)” is the joint exceedance probability. “Heat ratio vs W”, “PM ratio vs W”, and “Joint ratio vs W” are the within-group exceedance rates divided by the corresponding predominantly White reference rates from Table 2 (P_LST=11.06%, P_PM=13.15%, P_Joint=3.33%). “Mixed” denotes tracts in which no single racial group constitutes 50% or more of the population. Asian-predominant tracts are tracts in which non-Hispanic Asian residents constitute 50% or more of the population. These two groups account for 8,723 of the 42,304 analytic-sample tracts; the remaining 33,581 are reported by predominant racial group in Table 2 of the main manuscript.*

**Table S10. The 48 metropolitan areas in the analytic sample**

*Core Based Statistical Areas (CBSAs) in the analytic sample, with CBSA Federal Information Processing System (FIPS) codes from the 2020 Office of Management and Budget delineation. The 48 CBSAs cover approximately 174.6 million residents (52% of the US population). San Juan-Bayamón-Caguas, PR is the only CBSA in the integrated exposure database not included; it is excluded because the EPA Fused Air Quality Surface Downscaler used for PM2.5 does not cover Puerto Rico.*

| **#** | **CBSA code** | **CBSA name** |
| --- | --- | --- |
| 1 | 12060 | Atlanta-Sandy Springs-Roswell, GA |
| 2 | 12420 | Austin-Round Rock-San Marcos, TX |
| 3 | 12580 | Baltimore-Columbia-Towson, MD |
| 4 | 13820 | Birmingham, AL |
| 5 | 14460 | Boston-Cambridge-Newton, MA-NH |
| 6 | 15380 | Buffalo-Cheektowaga, NY |
| 7 | 16740 | Charlotte-Concord-Gastonia, NC-SC |
| 8 | 16980 | Chicago-Naperville-Elgin, IL-IN |
| 9 | 17140 | Cincinnati, OH-KY-IN |
| 10 | 18140 | Columbus, OH |
| 11 | 19100 | Dallas-Fort Worth-Arlington, TX |
| 12 | 19740 | Denver-Aurora-Centennial, CO |
| 13 | 19820 | Detroit-Warren-Dearborn, MI |
| 14 | 26420 | Houston-Pasadena-The Woodlands, TX |
| 15 | 26900 | Indianapolis-Carmel-Greenwood, IN |
| 16 | 27260 | Jacksonville, FL |
| 17 | 28140 | Kansas City, MO-KS |
| 18 | 29820 | Las Vegas-Henderson-North Las Vegas, NV |
| 19 | 31080 | Los Angeles-Long Beach-Anaheim, CA |
| 20 | 31140 | Louisville/Jefferson County, KY-IN |
| 21 | 32820 | Memphis, TN-MS-AR |
| 22 | 33100 | Miami-Fort Lauderdale-West Palm Beach, FL |
| 23 | 33340 | Milwaukee-Waukesha, WI |
| 24 | 33460 | Minneapolis-St. Paul-Bloomington, MN-WI |
| 25 | 34980 | Nashville-Davidson--Murfreesboro--Franklin, TN |
| 26 | 35380 | New Orleans-Metairie, LA |
| 27 | 35620 | New York-Newark-Jersey City, NY-NJ |
| 28 | 36420 | Oklahoma City, OK |
| 29 | 36740 | Orlando-Kissimmee-Sanford, FL |
| 30 | 37980 | Philadelphia-Camden-Wilmington, PA-NJ-DE-MD |
| 31 | 38060 | Phoenix-Mesa-Chandler, AZ |
| 32 | 38300 | Pittsburgh, PA |
| 33 | 38900 | Portland-Vancouver-Hillsboro, OR-WA |
| 34 | 39300 | Providence-Warwick, RI-MA |
| 35 | 39580 | Raleigh-Cary, NC |
| 36 | 40060 | Richmond, VA |
| 37 | 40140 | Riverside-San Bernardino-Ontario, CA |
| 38 | 40900 | Sacramento-Roseville-Folsom, CA |
| 39 | 41180 | St. Louis, MO-IL |
| 40 | 41620 | Salt Lake City-Murray, UT |
| 41 | 41700 | San Antonio-New Braunfels, TX |
| 42 | 41740 | San Diego-Chula Vista-Carlsbad, CA |
| 43 | 41860 | San Francisco-Oakland-Fremont, CA |
| 44 | 41940 | San Jose-Sunnyvale-Santa Clara, CA |
| 45 | 42660 | Seattle-Tacoma-Bellevue, WA |
| 46 | 45300 | Tampa-St. Petersburg-Clearwater, FL |
| 47 | 47260 | Virginia Beach-Chesapeake-Norfolk, VA-NC |
| 48 | 47900 | Washington-Arlington-Alexandria, DC-VA-MD-WV |

*Note: CBSA codes are 5-digit FIPS codes from the OMB CBSA delineation. CBSAs are sorted alphabetically by name. Total population across the 48 CBSAs is approximately 174.6 million (ACS 2016 to 2020 five-year estimates), or roughly 52% of the US population. The 48 represent essentially all of the most populous large US metropolitan areas in the contiguous United States with complete tract-level coverage in our integrated exposure database (FAQSD for PM2.5 and ozone, MODIS LST for surface temperature).*

**Supplementary methodological notes**

**S.M.1 Bootstrap procedure for CER intervals**

The within-group conditional exceedance ratio at the 80th percentile (CER80) is defined in the main text Eq. 3 as the ratio of the observed joint exceedance rate to the rate expected under statistical independence of the two stressors within the predominant-racial-composition subsample. To respect within-metropolitan-area spatial dependence, we constructed confidence intervals using a CBSA-block bootstrap that resamples CBSAs with replacement and retains all tracts within each resampled CBSA.

The 80th-percentile thresholds for LST and PM2.5 exceedance were computed once on the full 48-CBSA analytic sample, within each CBSA, and held fixed across all bootstrap replicates and jackknife folds. This anchoring is essential: in an early iteration of the analysis we recomputed thresholds within each resampled CBSA set, which produced a bootstrap distribution that drifted systematically away from the full-sample point estimate (by 19% in the White-predominant group). Fixing the thresholds at the full-sample within-CBSA q80 anchors the bootstrap distribution on the full-sample estimand and recovers BCa intervals that contain their point estimates.

BCa intervals (Efron 1987) were computed from 5,000 bootstrap replicates with the acceleration parameter estimated from a leave-one-CBSA-out cluster jackknife using the same CER estimator. The bias-correction parameter z0 was estimated from the proportion of bootstrap replicates falling below the full-sample point estimate, with the proportion bounded away from zero and one to avoid the BCa formula breaking at the tails. Under this construction, the bootstrap means closely approximate the full-sample point estimates (z0 ≈ 0 for all three groups, indicating well-centred resampling), and the BCa intervals contain their point estimates for all three groups.

**S.M.2 Spatial autoregressive sensitivity**

As a sensitivity check on the multilevel regression coefficient on percent people of colour, we fitted spatial autoregressive (SAR) lag models in the 10 largest CBSAs separately. In each CBSA, the SAR specification was Y = ρWY + Xβ + ε, where W is the row-standardised queen-contiguity weights matrix and ρ is the spatial autoregressive parameter. The mean within-CBSA POC coefficient declined from 0.366 in the Model 4 multilevel specification to approximately 0.04 in the SAR specification. We interpret the multilevel coefficient as a population-average gradient that absorbs spatial co-clustering of demographic and environmental features, and the SAR estimate as a stricter conditional-association estimate that is, by construction, harder to identify in spatially structured data. The descriptive joint disparity (Section 3.2), the CER analysis (Section 3.5), and the HOLC analysis (Section 3.3) are computed outside the multilevel-regression framework and are unaffected by the SAR specification.

**S.M.3 Multiple-testing correction**

We computed Benjamini-Hochberg and Bonferroni adjustments across the six primary hypothesis tests (POC β in CCEI multilevel model; POC OR in joint hotspot logistic; POC RR in joint hotspot Poisson; HOLC D-vs-A OR; White CER80 above 1; Hispanic CFG λU below White CFG λU). All six survive both corrections at the 0.05 level.
